# Modelling the impact of changes to prescription medicine cost-sharing schemes among middle aged and older adults

**DOI:** 10.1101/2025.01.21.25320791

**Authors:** James Larkin, Ciaran Prendergast, Logan T. Murry, Michelle Flood, Barbara Clyne, Sara Burke, Conor Keegan, Fiona Boland, Tom Fahey, Nav Persaud, Rose Anne Kenny, Frank Moriarty

**Affiliations:** School of Pharmacy and Biomolecular Sciences, RCSI University of Medicine and Health Sciences, Dublin, Ireland; Department of General Practice, RCSI University of Medicine and Health Sciences, Dublin, Ireland; Department of Epidemiology and Public Health, School of Population Health, RCSI University of Medicine and Health Sciences, Dublin, Ireland; Centre for Health Policy and Management, School of Medicine, Trinity College Dublin, Dublin, Ireland; Economic and Social Research Institute, Dublin, Ireland; Data Science Centre, School of Population Health, RCSI University of Medicine and Health Sciences, Dublin, Ireland; Department of Family and Community Medicine, University of Toronto, Toronto, Ontario, Canada; MAP Centre for Urban Health Solutions, St Michael’s Hospital, Toronto, Ontario, Canada; The Irish Longitudinal Study on Ageing, Trinity College Dublin, Dublin, Ireland

## Abstract

**Objective:** To assess impacts of government changes to prescription medicine co-payments on individuals’ out-of-pocket prescription medicine expenditure.

**Methods:** Data from The Irish Longitudinal Study on Ageing were used. Participants were community-dwelling adults aged ≥56 years. Ireland has two prescription cost-sharing schemes: the General Medical Services (GMS) scheme (primarily for low-income populations), involving a low monthly payment cap and low co-payments, and the Drugs Payment Scheme (DPS) (for others), with a higher cap and no co-payment limit. We modelled changes to these schemes implemented between 2016-2022 using 2016 data, assessing impacts on out-of-pocket prescription medicine expenditure across participant characteristics.

**Results:** Among 4,155 participants with out-of-pocket prescription medicine expenditure, estimated mean annual prescription medicine expenditure for GMS-eligible participants reduced from €117 (95%CI=€114-120) to €55 (95%CI=€54-€56) due to post-2016 scheme changes. For DPS-eligible participants it reduced from €719 (95%CI=€694-€744) to €555 (95%CI=€541-€569). Those on more medicines had greater savings, with similar savings across income groups.

**Conclusions:** Co-payment changes led to average savings of €62 for GMS-eligible participants and €174 for DPS-eligible participants. Although absolute savings were smaller for GMS participants, these were likely more impactful for this low-income population. Further reductions in monthly caps and co-payment charges, particularly for low-income populations, warrant consideration.

## 1. Background

Prescription medicine use has increased across countries^1–6^ and concurrently, there has been increased development of expensive medicines,^7^ leading to increased medicine expenditure for countries and individuals. Medicines represent a significant proportion of out-of-pocket healthcare costs in Ireland and other countries,^8–10^ particularly for low-income groups.^11^ National prescription cost-sharing policies vary, reflecting differences in public financing and health insurance policies. Prescription medicines co-payment levels increased between 2009-2016 in Ireland following the financial crisis, due to changes to state-provided schemes.^12^ Since 2016, however, co-payment levels were reduced due to further schemes changes,^13,14^ with more changes under consideration.^15–18^

Increased out-of-pocket prescription medicine expenditure, linked to increased co-payment charges, can lead to cost-related non-adherence (even if changes are small),^19^ with negative impacts on health.^20^ Greater out-of-pocket healthcare expenditure can also force people to cut spending on essentials like food,^21^ further affecting wellbeing.^21^

### 1.1 Prescription Cost-Sharing and Reform in Ireland

In Ireland, low-income individuals are eligible for the General Medical Services (GMS) scheme, which includes a reduced co-payment: €2.50 per prescription item in 2016, capped at €25 monthly for a household. Income eligibility for this scheme depends on age and household size, with higher thresholds for older adults.^22^ Those without GMS scheme eligibility are eligible for the Drugs Payment Scheme (DPS), where they pay full price for each medicine up to a monthly household cap (which peaked at €144 in 2016) with the remaining costs covered by the state.^23^ In 2016, 36% of the population were GMS-eligible and the remainder were DPS-eligible.^24^ Additionally, those with 16 specific conditions (e.g. diabetes, Parkinson’s) are eligible for the Long-Term Illness (LTI) scheme which provides for prescription medicines prescribed for that condition at no charge.^25^ Further details of cost-sharing schemes are in eBox 1.

In May 2017, the Sláintecare Report from an all-parliamentary committee on Irish healthcare reform recommended large increases in public healthcare funding.^18^ One of its goals was to reduce out-of-pocket prescription medicine costs for individuals.^18^ Following this report, prescription co-payment charges and monthly payment caps have been significantly reduced (as described in Table 1), but there has been limited assessment of the financial impact of these changes on individuals. Whilst Ireland’s healthcare coverage system is relatively unique, this study is relevant for other countries facing similar challenges in providing affordable healthcare.^26,27^

**Table 1.** Prescription cost-sharing scheme changes affecting co-payments and monthly caps modelled for analysis.

| **Scheme Change Number** | **General Medical Services Scheme prescription medicine cover** | **Drugs Payment Scheme prescription medicine upper monthly cap** |
| --- | --- | --- |
| Baseline | 01 December 2013: €2.50 per item and a €25 upper monthly cap<sup>34</sup> | 1 January 2013: €144<sup>35</sup> |
| Change 1 | 1 January 2018: €2 per item and a €20 upper monthly cap<sup>36</sup> | 1 January 2018: €134<sup>37</sup> |
| Change 2 | 1 April 2019: Aged 70 and over: €1.50 per item and a €15 upper monthly cap<br>Aged under 70: €2 per item and a €20 upper monthly cap<sup>38</sup> | 1 April 2019: €124<sup>37</sup> |
| Change 3 | 01 November 2020: Aged 70 and over: €1 per item and a €10 upper monthly cap<br>Aged under 70: €1.50 per item and a €15 upper monthly cap<sup>39</sup> | 01 November 2020: €114<sup>37</sup> |
| Change 4 | No change (same as *Change 3*) | 1 January 2022: €100<sup>37</sup> |
| Change 5 | No change (same as *Change 3*) | 1 March 2022: €80<sup>37</sup> |

### 1.2 Aim

This study aims to assess the impact of changes to prescription medicine co-payments and monthly payment caps on individuals’ out-of-pocket prescription medicine expenditure.

## 2. Methods

This is a modelling study using Wave 4 data from The Irish Longitudinal Study on Ageing (TILDA), collected in 2016.^28^ TILDA is a nationally representative prospective cohort study charting the health, economic, and social circumstances of adults in Ireland aged ≥56 years. Wave 4 was the most recent data available where prescription medicine co-payment charges and caps were at their highest level since 2004.^29^ Between 2016-2022, changes to prescription medicine charges and monthly payment caps were implemented (see Table 1).

### 2.2 Participants

At baseline (2009), recruited participants were ≥50 years. Spouses/partners of recruited participants were also recruited, regardless of age. Households were randomly selected using a national geodirectory. For this analysis, participants were those at Wave 4 (then aged ≥56 years) free from significant cognitive impairment. We excluded a) those in residential care settings because their healthcare utilisation patterns likely differ from community-dwelling adults, and b) participants who had not answered the out-of-pocket prescription medicine expenditure question.

### 2.3 Variables and data sources

Out-of-pocket prescription medicine expenditure information was gathered with the question: “*Not counting health insurance refunds, on average about how much do you pay out-of-pocket for your prescribed drugs per month*?” (see eBox 2). We multiplied this value by 12 to estimate annual expenditure. If reported monthly expenditure was >€288 (twice the DPS monthly cap) it was excluded from the analysis. Exceptions were provided for if there were clear errors (e.g. decimal point error), as detailed in eTable 1. Neither the question nor the analysis accounted for tax relief on out-of-pocket prescription medicine expenditure (see eBox 1 for more details of tax relief policies).

Healthcare eligibility details were gathered with questions on Ireland’s publicly-funded schemes and private health insurance (see eBox 3). Health conditions were identified by asking about doctor-diagnosed conditions (see eBox 4). For the condition count, we used TILDA’s list of 36 conditions and combined some conditions to develop 21 broader conditions (eBox 5) in line with previous TILDA studies.^30,31^ In the descriptive analysis, we categorised the number of chronic conditions as 0, 1, 2, or 3+. We also included a complex multimorbidity variable; defined as an individual having ≥3 chronic conditions with at least three conditions each primarily affecting one distinct body systems (see eBox 6).^32^ Participants also reported ‘regular’ medicines which they took ‘every day or every week’ (details in eBox 2). The count for this variable was split into quintiles for analysis.

Demographic variables included age, sex, residence (urban or not urban), marital status, equivalised household income (see eBox 7) and educational attainment. Equivalised household income was split into quintiles for analysis, based on the entire sample.

### 2.4 Scheme changes modelled and cost estimation

Several scheme changes affecting co-payments and monthly payment caps were modelled using varying GMS prescription charges, GMS payment caps and DPS payment caps (Table 1). These changes were introduced post-2016. The models assume scheme changes were applied in 2016, and do not reflect changes in prescribed medicines that may have occurred over time or inflation in costs of medicines.

To calculate costs for participants covered by the GMS scheme, the number of potentially reimbursable medicines was determined.^33^ Costs were calculated by multiplying the number of relevant medicines by the prescription charge, up to the relevant monthly cap (Table 1). For participants without GMS cover, their reported monthly out-of-pocket prescription medicine expenditure was truncated at the relevant DPS cap (Table 1), where necessary. For those covered by the LTI scheme, medicines relating to conditions on the LTI scheme were excluded from calculations of co-payment charges for the GMS scheme and number of prescribed medicines used as a covariate.

### 2.5 Data analysis

For each scheme change, descriptive statistics were generated to estimate annual out-of-pocket prescription medicine expenditure across demographics, government prescription cost-sharing schemes and health characteristics. Bar charts were generated depicting mean savings associated with each co-payment/cap change. Dot plots were generated depicting relative reduction in expenditure (as a percentage) associated with each change to co-payments and monthly caps. Expenditure analysis and modelling was only conducted for those with any out-of-pocket prescription medicine expenditure, those who reported no expenditure on out-of-pocket on prescription medicines were excluded.

## 3. Results

There were 5,912 participants. After exclusion of 78 participants in residential care, 153 who did not report out-of-pocket healthcare expenditure and 13 whose out-of-pocket prescription medicine expenditure was deemed to be an invalid outlier, there were 5,668 eligible participants. Demographic and out-of-pocket prescription medicine expenditure details for that group are in eTable 2. Of the 5,668, 73.3% (n=4,155) had out-of-pocket prescription medicine expenditure. Of those, 56.5% (n=2,348) were female, and 75.6% (n=3,143) took ≥2 regular medicines. Table 2 describes their demographics, cost-sharing scheme coverage, health characteristics and expenditure.

**Table 2.** Out-of-pocket (OOP) prescription medicine expenditure at Wave 4 before scheme changes, among those with expenditure, by participant characteristics (n=4,155)

|  | General Medical Services Scheme Eligible (N=2,214) |  |  | Drugs Payment Scheme eligible (N=1,941)<sup>i</sup> |  |  |
| --- | --- | --- | --- | --- | --- | --- |
|  | Total % (n) | Mean OOP Prescription Medicine Expenditure (SD) | Median OOP Prescription Medicine Expenditure (IQR) | Total % (n) | Mean OOP Prescription Medicine Expenditure (SD) | Median OOP Prescription Medicine Expenditure (IQR) |
| **Overall** | 100.0% (2,214) | €117 (€76) | €90 (60-150) | 100.0% (1,941) | €719 (€571) | €480 (€240-1,140) |
| **Age (years)** |  |  |  |  |  |  |
| <60 | 9.1% (202) | €97 (€71) | €90 (30-150) | 19.8% (384) | €624 (€542) | €420 (€216-840) |
| 60-69 | 28.8% (638) | €107 (€71) | €90 (60-150) | 49.6% (962) | €678 (€548) | €480 (€240-960) |
| 70-79 | 39.6% (877) | €120 (€78) | €120 (60-180) | 23.4% (455) | €788 (€601) | €600 (€300-1,440) |
| 80+ | 22.4% (497) | €135 (€78) | €120 (90-180) | 7.2% (140) | €1,034 (€579) | €1,044 (€480-1,728) |
| **Sex** |  |  |  |  |  |  |
| Female | 58.8% (1,302) | €121 (€77) | €120 (60-180) | 53.9% (1046) | €708 (€564) | €480 (€240-1,080) |
| Male | 41.2% (912) | €112 (€75) | €90 (60-150) | 46.1% (895) | €732 (€580) | €480 (€240-1,200) |
| **Education** |  |  |  |  |  |  |
| Primary/none | 40.9% (905) | €125 (€79) | €120 (60-180) | 10.9% (211) | €744 (€560) | €576 (€300-1,140) |
| Secondary | 39.8% (882) | €114 (€75) | €90 (60-150) | 37.5% (727) | €730 (€578) | €480 (€264-1,200) |
| Third/higher | 19.3% (427) | €109 (€71) | €90 (60-150) | 51.7% (1003) | €706 (€569) | €480 (€240-1,080) |
| **Equivalised annual household income (quintiles)\*** |  |  |  |  |  |  |
| <€6,000 | 35.6% (700) | €122 (€78) | €120 (60-180) | 9.1% (153) | €698 (€575) | €456 (€216-1,080) |
| €6,006-€10,000 | 25.7% (507) | €115 (€76) | €90 (60-150) | 13.1% (222) | €671 (€538) | €480 (€240-960) |
| €10,029-€15,133 | 21.1% (416) | €120 (€77) | €120 (60-150) | 19.7% (333) | €778 (€596) | €576 (€300-1,320) |
| €15,142-€24,000 | 12.5% (246) | €111 (€75) | €90 (60-150) | 27.0% (457) | €734 (€575) | €540 (€264-1,104) |
| >€24,000 | 5.1% (100) | €104 (€65) | €90 (60-150) | 31.1% (525) | €703 (€565) | €480 (€240-1,020) |
| **Area of residence\*** |  |  |  |  |  |  |
| Urban | 47.3% (1,048) | €121 (€79) | €120 (60-180) | 56.7% (1100) | €763 (€591) | €540 (€276-1,200) |
| Not Urban | 52.7% (1,166) | €114 (€73) | €90 (60-150) | 43.3% (841) | €661 (€539) | €480 (€240-912) |
| **Marital status** |  |  |  |  |  |  |
| Partnered | 58.4% (1,294) | €112 (€73) | €90 (60-150) | 77.1% (1496) | €733 (€582) | €522 (€240-1,200) |
| Not Partnered | 41.6% (920) | €124 (€80) | €120 (60-180) | 22.9% (445) | €672 (€529) | €480 (€270-960) |
| **Private health insurance** |  |  |  |  |  |  |
| Yes | 36.1% (800) | €115 (€71) | €90 (60-150) | 85.3% (1656) | €731 (€576) | €504 (€240-1,200) |
| No | 63.9% (1,414) | €119 (€79) | €120 (60-180) | 14.7% (285) | €648 (€535) | €432 (€240-840) |
| **General Practitioner Visit Card** |  |  |  |  |  |  |
| Yes | - | - | - | 26.0% (505) | €854 (€601) | €720 (€312-1,560) |
| No | 100.0% (2,214) | €117 (€76) | €90 (60-150) | 74.0% (1,436) | €671 (€552) | €480 (€240-960) |
| **Long term illness (LTI) scheme** |  |  |  |  |  |  |
| Yes | 9.0% (199) | €96 (€77) | €90 (30-150) | 5.4% (104) | €868 (€616) | €720 (€336-1,716) |
| No | 91.0% (2,015) | €119 (€76) | €120 (60-180) | 94.6% (1837) | €711 (€567) | €480 (€240-1,080) |
| **Number of prescription medicines (quintiles)\*** |  |  |  |  |  |  |
| 0 regular medicines** | 4.8% (106) | €0 (€0) | €0 (0-0) | 4.1% (79) | €444 (€420) | €360 (€120-576) |
| 1 regular medicine | 13.4% (297) | €30 (€0) | €30 (30-30) | 27.3% (530) | €366 (€404) | €240 (€132-384) |
| 2-3 regular medicines | 31.8% (705) | €75 (€15) | €60 (60-90) | 39.4% (765) | €655 (€488) | €480 (€300-840) |
| 4-5 regular medicines | 25.3% (560) | €134 (€15) | €120 (120-150) | 18.4% (358) | €1,056 (€529) | €960 (€600-1,728) |
| 6+ regular medicines | 24.7% (546) | €225 (€44) | €210 (180-270) | 10.8% (209) | €1374 (€467) | €1,692 (€1,080-1,728) |
| **Number of chronic conditions** |  |  |  |  |  |  |
| 0 chronic conditions | 6.4% (141) | €83 (€61) | €60 (30-120) | 8.3% (162) | €545 (€513) | €360 (€192-600) |
| 1 chronic condition | 18.4% (408) | €86 (€61) | €60 (30-120) | 28.5% (554) | €532 (€506) | €330 (€180-696) |
| 2 chronic conditions | 24.6% (545) | €105 (€64) | €90 (60-150) | 29.4% (570) | €710 (€543) | €480 (€288-960) |
| 3+ chronic conditions | 50.6% (1,120) | €139 (€81) | €120 (90-195) | 33.7% (655) | €928 (€590) | €780 (€360-1,680) |
| **Complex multimorbidity** |  |  |  |  |  |  |
| Yes | 43.5% (963) | €142 (€82) | €150 (90-210) | 28.0% (544) | €951 (€578) | €840 (€420-1,680) |
| No | 56.5% (1,251) | €98 (€65) | €90 (60-150) | 72.0% (1,397) | €629 (€542) | €420 (€240-840) |

<sup>f</sup>includes all those without GMS scheme cover who report any out-of-pocket prescription medicine expenditure.
\*This variable had data missing and the relevant percentages are calculated with missing data excluded.
\* Excluded medicines not covered by state drug schemes as these are primarily over-the-counter medicines and supplements. Also excluded medicines always indicated for a condition covered by the LTI scheme.
\*\* Though these participants are on 0 regular medicines, the expenditure question does not preclude participants from including expenditure on non-regular medicines
Note: SD=Standard Deviation. IQR=Interquartile Range. GMS=General Medical Services. DPS=Drugs Payment Scheme

Mean annual out-of-pocket prescription medicine expenditure for those with DPS cover, was €719 (SD=€571) when the €144 monthly cap was in place, which then decreased to €555 (SD=€325) when modelling the €80 monthly cap, i.e. mean annual savings of €164 (SD=€296). Mean savings associated with each DPS change are in Figure 1. The largest mean savings occur for those on ≥6 regular medicines (€490, SD=€331), with 36.5% (mean=€179, SD=€102) of savings resulting from the €100 to €80 change in cap. For those aged ≥80 years, mean savings were €292 (SD=€340) with 39.9% (mean=€117, SD=€118) of savings resulting from the €100 to €80 change in cap. Details of percentage savings are in eFigure 1. eTable 3 details the number and percentage of people whose out-of-pocket prescription medicine expenditure is affected by DPS monthly cap changes.

**Figure 1.**
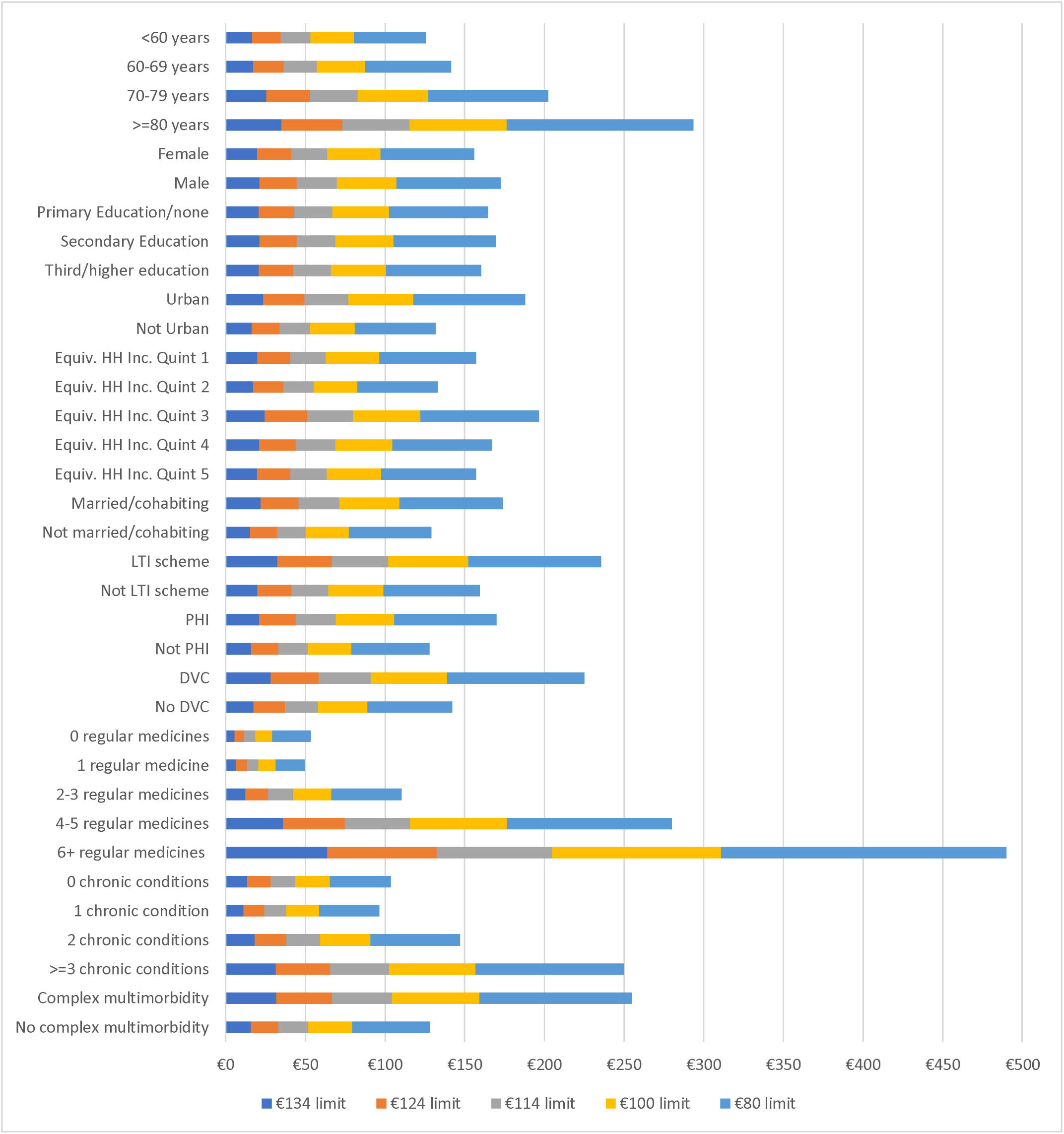
Mean savings associated with each change to Drugs Payment Scheme monthly caps Note: Equiv. HH Inc. Quint=Equivalised Household Income Quintile. DVC=General Practitioner Visit Card. LTI=Long Term Illness. PHI=Private Health insurance.

Mean savings associated with each GMS scheme change are in Figure 2. Mean expenditure for those with GMS scheme cover was €117 (SD=€76) at baseline, which decreased to €55 (SD=€36) under change 3, i.e. a mean annual savings of €62 (SD=€44). The largest savings occur for those on ≥6 regular medicines (mean=€122, SD=€32), with 36.6% (mean=€45, SD=€9) of these savings occurring due to change 3. For those aged ≥80 years, their mean savings were €81 (SD=€47) with 33.3% (mean=€37, SD=€16) of savings resulting from change 3. eFigure 2 provides further details of percentage savings. eTable 4 details the number and percentage of people whose annual out-of-pocket prescription medicine expenditure is affected by changes to the GMS scheme monthly cap and prescription charges.

**Figure 2.**
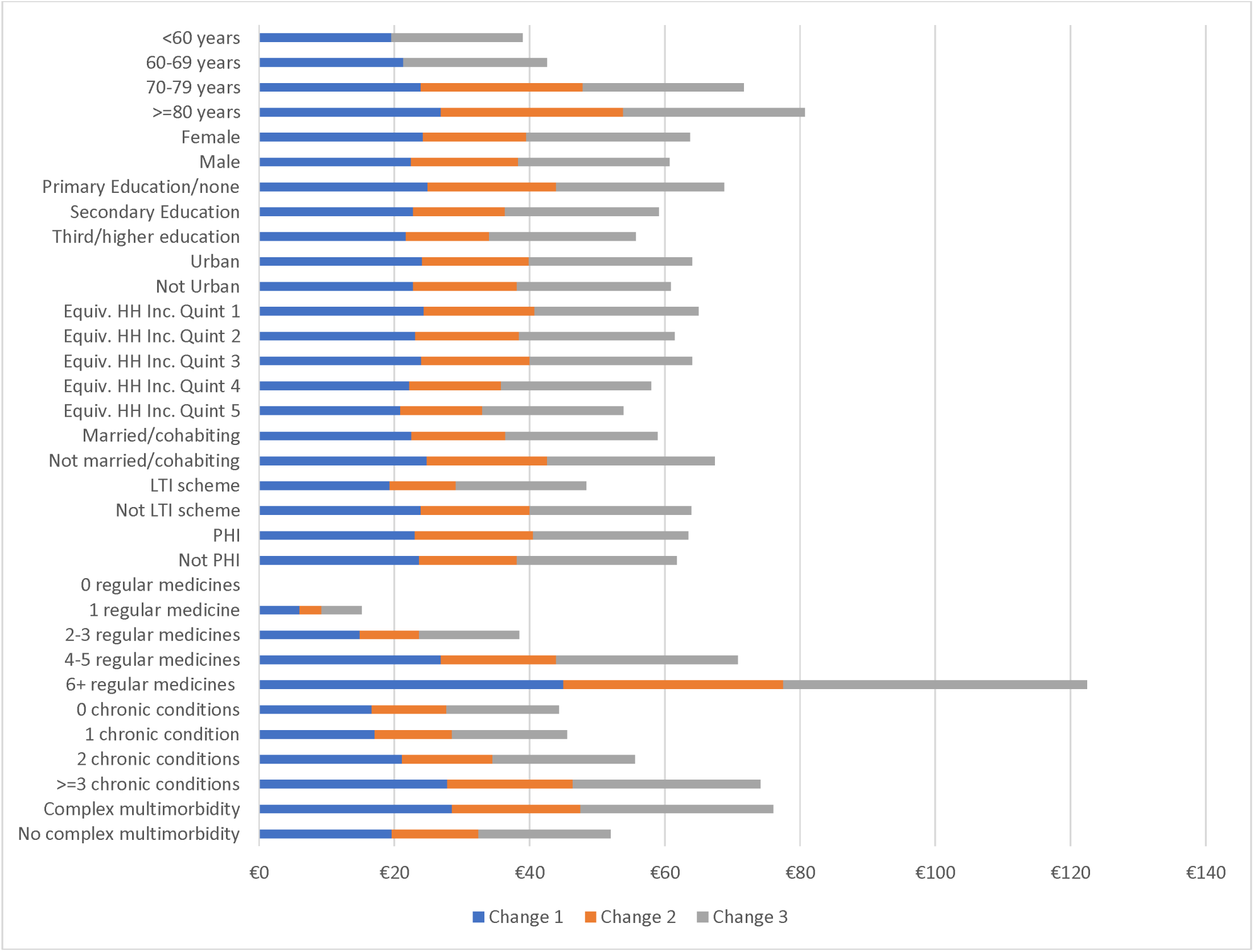
Mean savings associated with each GMS change Note: GMS=General Medical Services. Equiv. HH Inc. Quint=Equivalised Household Income Quintile. DVC=General Practitioner Visit Card. LTI=Long Term Illness. PHI=Private Health insurance.

## 4. Discussion

### 4.1 Summary

The changes to co-payment charges and caps for prescription medicines in Ireland since 2016 likely led to a number of reductions in out-of-pocket prescription medicine expenditure. The effect is strong for those with DPS cover: reducing the monthly payment cap from €144 to €80 is estimated to have reduced average expenditure from €719 to €555, translating to mean annual savings of €164. Mean reductions ranged from €50 to €490 across demographic groups, with reductions necessarily concentrated amongst higher spenders.

For GMS cover, estimated reductions were smaller in absolute terms, with mean expenditure reducing from €117 to €55, an estimated mean annual savings of €62. However, as this group has lower incomes, these savings may be more impactful. Estimated mean reductions ranged from €10 to €123 across demographic groups.

Estimated savings across income quintiles were similar in both schemes, despite potentially greater need for reductions among lower income groups, especially those in the GMS scheme, where small co-payment changes affect cost-related non-adherence.^40^ It is important to note that the two approaches to co-payment reductions differ: for the GMS scheme both the monthly payment cap and co-payment charges were reduced, while for the DPS, only the monthly payment cap was lowered, individual co-payment charges are not capped. In both groups the estimated savings were highest among those on six or more regular medicines.

### 4.2 Implications

There is still significant potential to reduce prescription co-payments. For example, Northern Ireland, has no prescription medicine co-payments.^41^ Further reductions should prioritise expanding GMS eligibility by raising the income threshold for eligibility to include those DPS-eligible individuals with low incomes. Though this may lead to significantly increased healthcare utilisation.^42^ Reductions should also prioritise those with GMS eligibility as they have lower capacity to pay, even for small co-payment charges.^12^ Additional strategies, such as deprescribing interventions to reduce the number of regular medicines safely,^43^ could further alleviate the prescription cost burden. Future research could model possible changes to prescription cost-sharing schemes including those proposed by political parties^15–18^ or policies in other countries. This could also involve modelling the potential administrative savings associated with simplified universal entitlements.^44^

### 4.3 Strengths and limitation

A strength of this study is the nationally representative sample captured by TILDA. In terms of limitations, use of self-report may reduce accuracy for some complex variables such as income, however there is some evidence on the accuracy of self-reported medicine use.^45,46^ The primary limitation of this study is that payment levels and savings are estimated, and estimates after 2016 do not account for inflation or changes in the population’s prescription medicine use and income levels. Finally, using reported monthly expenditure to estimate annual expenditure may not have accurately captured annual expenditure (i.e. omitting once-off acute medicine), though the shorter recall period likely reduces recall bias.^47^

### 4.4 Conclusion

Changes to prescription medicine co-payments and caps under various government schemes in Ireland have likely led to large savings on out-of-pocket prescription medicine expenditure. These savings were estimated to be higher in absolute terms for those with DPS cover but were perhaps more impactful for those with GMS scheme cover given their lower incomes. There is still a high prescription medicine cost burden for individuals in Ireland, compared to other European countries, and consideration could be given to further reductions in monthly payment caps and co-payment charges, particularly for those with low incomes.

## Data Availability

Researchers interested in using TILDA data can find out details about how to access it here: https://tilda.tcd.ie/data/accessing-data/

