## Appendix A for "Modelling the impact of changes to prescription medicine cost-sharing schemes among middle aged and older adults"

### eBox 1. Additional information about cost-sharing schemes in Ireland

The *GP Visit Card* provides for no charge general practitioner (GP) visits but no additional prescription medicine entitlements.^1^ The Health Amendment Act (HAA) provides those who were affected by State provided contaminated blood products to coverage similar to the GMS scheme.^2^ To simplify analysis, the HAA card was grouped with the GMS scheme.

An individual can claim income tax relief on out-of-pocket health expenditure (including on prescription medicines) at the rate of 20% of their out-of-pocket health expenditure.^3^ In a single year, the maximum amount of out-of-pocket health expenditure an individual can claim on is €1,000, which would mean €200 in tax relief.^3^

### eBox 2. Medicine questions in TILDA wave 4

**HU035: Not counting health insurance refunds, on average about how much [do/does] [you/he/she] pay out-of-pocket for [your/his/her] prescribed drugs per month?**

IWER: IF RESPONDENT DOES NOT PURCHASE PRESCRIBED DRUGS REGULARLY, ASK FOR TOTAL SPENT IN THE LAST 12 MONTHS IN PRESCRIBED DRUGS AND DIVIDE BY 12.

IWER: IF R CANNOT GIVE EXACT VALUE, ACCEPT APPROXIMATE VALUE

VALUE [Allow Responses up to 2 decimal places]

€0.00 … €5,000.00

-98. DK

-99. RF

NOTE: Include the €2.50 per prescribed item charges for medical card holders.

NOTE: Do not consider expenses for self-medication or drugs not prescribed

NOTE: By ‘out of pocket’ expenses we mean everything that is not paid by the insurance company. If [you/he/she] first [pay/pays] but later get it refunded, this is not out of pocket expenses.

(SHARE)

**MD001a:** **Now I would like to record all medications that [you/Rname] [take/takes] on a regular basis, like every day or every week. This will include prescription and non-prescription medications, over-the-counter medicines, vitamins, and herbal and alternative medicines.**

**Do/Does [you/Rname] take any medication on a regular basis?**

1. Yes	**GO TO MD001**

5. No

98. DK

99. RF

**IF INTSTATUSW4=1,2 OR 3 & MD001A = 5 GO TO MD008**

**IF INTSTATUSW4= 4 OR 5 & MD001A = 5 GO TO NEXT SECTION**

**IWER: ASK RESPONDENT IF YOU COULD SEE THE MEDICATIONS HE/SHE TAKES SO YOU CAN COPY DOWN THE CORRECT SPELLING OF EACH TABLET.**

**IWER: PROMPT: DO I HAVE ALL OF [YOUR/RName’s] MEDICATIONS HERE?**

**DISPLAY NOTE: INPUT THE BRAND NAME WHEN AVAILABLE, RATHER THAN THE GENERIC NAME FOR ANY MEDICATIONS**

**MD001: TYPE THE FIRST FOUR LETTERS OF THE MEDICATION. YOU WILL BE GIVEN A LIST OF POSSIBILITIES CHOOSE ONE. Each medication is recorded in a separate variable**

**(MD001_01 – MD001_20; MD001_ATC_01 - MD001_ATC_20; MD001_NONPROP_01 - MD001_NONPROP_20)**

95. Other (specify) GO TO MD001oth

98. DK

99. RF

**MD001oth: Other (specify)**

**IF THE MEDICATION DOES NOT APPEAR ON THE LIST CAREFULLY TYPE THE FULL MEDICATION NAME. MAKE SURE YOU TYPE THE NAME OF THE BRAND USED AND NOT THE CHEMICAL NAME.**

**(MD001OTH_01-MD001OTH_20)**

Text: up to 60 characters GO TO MD005

**MD005: Was this medication prescribed by a doctor or did you get it over the counter? (MD005_01-MD005_20)**

1. Prescribed by a doctor

2. Over the counter

98. DK

99. RF

**(Note to Scripters -** For medications that are available over the counter as well as on prescription, we are interested in how the respondent got them.)

**RETURN AND REPEAT MD001 FOR UP TO 20 MEDICATIONS PER PERSON**

**IF instatusW4= 4 OR 5 GO TO NEXT SECTION**

### eBox 3. Health coverage questions

**HU001: [Is/Are] [you/he/she] covered by:**

IWER: CODE THE ONE THAT APPLIES

1. Full Medical Card or equivalent

2. GP Visit Card

96. Neither of these

98. DK

99. RF

**Note:** This question is asked even of those covered by private medical insurance. Most over 70s are entitled to medical cards.

(EU-SILC)

**HU070: [Is/Are] [you/he/she] covered by:**

1. The long term illness scheme
2. A Health Act Amendment Card

96. Neither of these

98 DK

99 RF

**HU002: [Do/Does] [you/he/she] have private medical insurance cover (VHI etc.) in [your/his/her] own name or through another family member?**

1. Yes, in own name GO TO HU003

2. Yes, as the spouse of a subscriber GO TO HU003

3. Yes, as the relative of a subscriber GO TO HU003

5. No	GO TO HU049

98. DK	GO TO HU049

99. RF	GO TO HU049

(HEALTH INSURANCE AUTHORITY 2005 SURVEY)

### eBox 4. Question about health conditions in TILDA

(A) Since your last interview, has a doctor ever told you/ that you have any of the [other] conditions on this card?

(B) Has a doctor ever told [you/Rname] that [you/he/she] [have/has] any of the conditions on this card?

**IWER: PROBE - 'WHAT OTHERS?' CODE ALL THAT APPLY.**

1. Chronic lung disease such as chronic bronchitis
or emphysema	**GO TO PH302	[ph301_01]**

2. Asthma	**[ph301_02]**

3. Arthritis (including osteoarthritis, or rheumatism)	**GO TO PH304	[ph301_03]**

4. Osteoporosis, sometimes called thin or brittle bones	**[ph301_04]**

5. Cancer or a malignant tumour	**GO TO PH309	[ph301_05]**

(including leukaemia or lymphoma but excluding minor skin cancers)

6. Parkinson's disease	**GO TO PH314	[ph301_06]**

7. Any emotional, nervous or psychiatric problems,

such as depression or anxiety	**GO TO PH315	[ph301_07]**

9. Alzheimer's disease	**GO TO PH318	[ph301_09]**

10. Dementia, organic brain syndrome, senility	**GO** **TO PH319**	**[ph301_10]**

11. Serious memory impairment	**GO TO PH319a	[ph301_11]**

12. Stomach ulcers	**[ph301_12]**

13. Varicose Ulcers (an ulcer due to varicose veins)	**[ph301_13]**

14. Cirrhosis, or serious liver damage	**[ph301_14]**

15. Thyroid Problems	**GO TO PH325	[ph301_15]**

16. Alcohol abuse	**GO TO PH320a	[ph301_16]**

17. Substance abuse	**GO TO PH320b	[ph301_17]**

18. Chronic kidney disease	**GO TO PH327	[ph301_18]**

19. Severe Anaemia	**[ph301_19]**

20. Epilepsy	GO TO PH329	**[ph301_20]**

95. Other (please specify)	**[ph301_95] [ph301oth]**

96. None of these	**GO TO PH328	[ph301_96]**

98. DK	**GO TO PH328	[ph301_98]**

99. RF	**GO TO PH328	[ph301_99]**

(ELSA/ similar question HRS/NSHAP)

**IWER: SHOW CARD PH2 [PAGE #]**

**IF (intstatusW4 = 2, 4, 5), USE WORDING ‘B’. ALL OTHERS, USE WORDING ‘A’.**

PH201: Please look at card PH2.

(A) Since your last interview, has a doctor ever told you that you have any of the [other] conditions on this card?

(B) Has a doctor ever told [you/Rname] that [you/he/she] [have/has] any of the conditions on this card?

INTERVIEWER: PROBE - 'WHAT OTHERS?' CODE ALL THAT APPLY.

1. High blood pressure or hypertension	**[ph201_01]**

2. Angina	**[ph201_02]**

3. A heart attack

(including myocardial infarction or coronary thrombosis)	**[ph201_03]**

4. Congestive heart failure	**[ph201_04]**

5. Diabetes or high blood sugar	**[ph201_05]**

6. A stroke (cerebral vascular disease)	**[ph201_06]**

7. Ministroke or TIA	**[ph201_07]**

8. High cholesterol	**[ph201_08]**

9. A heart murmur	**[ph201_09]**

11. Atrial Fibrillation	**[ph201_11]**

12. An abnormal heart rhythm (not atrial fibrillation)	**[ph201_12]**

95. Any other heart trouble (please specify)	**[ph201_95][ph201oth]**

96. None of these	**[ph201_96]**

98. DK	**[ph201_98]**

99. RF	**[ph201_99]**

(ELSA/ similar questions in HRS/ SHARE)

(A) Since [your] last interview, has a doctor ever told you that you have any of the following [other] eye diseases?

(B) Has a doctor ever told [you/Rname] that [you/he/she] [have/has] any of the following eye diseases? [DISPLAY ALL CONDITIONS]

**IWER: READ OUT. CODE ALL THAT APPLY.**

1. Cataracts	**[ph105_01]**

2. Glaucoma	**[ph105_02]**

3. Age related macular degeneration	**[ph105_03]**

95. Other (please specify)	**[ph105_95] [ph105oth]**

### eBox 5. Conditions included in analysis (partly based on Ryan and colleagues)^4^

1. Cardiac Condition

- Angina
- Heart Attack
- Congestive Heart Failure
- Heart Murmur
- Abnormal heart rhythm
- Atrial fibrillation
- Any heart trouble

2. Cerebrovascular disease

- Stroke
- TIA

3. HTN
4. Diabetes
5. High Cholesterol
6. Chronic respiratory disease

- Chronic lung disease
- Asthma

7. Liver disease

- Alcohol abuse
- Cirrhosis
- Liver disease

8. Eye disease

- Cataracts
- Glaucoma
- Age related macular degeneration
- Other eye disease

9. Cognitive Impairment

- Alzheimer’s
  - Dementia
  - Serious cognitive impairment

10. Arthritis
11. Osteoporosis
12. Cancer
13. Parkinson’s disease
14. Emotional /psychological condition including anxiety and depression
15. Substance abuse
16. Stomach Ulcers
17. Varicose veins including varicose ulcers
18. Epilepsy
19. Thyroid Problems
20. Chronic kidney disease
21. Anaemia

### eBox 6. International Classification of Primary Care (ICPC) Classification of Body Systems

1. Cardiovascular
2. Respiratory
3. Digestive
4. Eyesystem
5. Psychological
6. Metabolic
7. Musculoskeletal
8. Neurological
9. Urological

### eBox 7. Equivalised Household Income details

Equivalised household income is an adjusted measure based on the OECD-modified equivalence scale for household size.^5^ It involves dividing household income by number of people in the household, with a weight of 1 applied to the first adult, 0.5 for each additional adult and 0.3 for each child.

### eTable 1. Prescription medicine expenditure outlier management

| Participant Response | Recoded to | N | Reason for recoding |
| --- | --- | --- | --- |
| €600 | €50 | 1 | Not a medical card holder and therefore, likely to be annual expenditure rather than monthly expenditure, therefore divided by 12 |
| €720 | €60 | 1 | Not a medical card holder and therefore, likely to be annual expenditure rather than monthly expenditure, therefore divided by 12 |
| €1032 | €86 | 1 | Not a medical card holder and therefore, likely to be annual expenditure rather than monthly expenditure, therefore divided by 12 |
| €1728 | €144 | 1 | Not a medical card holder and therefore, likely to be annual expenditure rather than monthly expenditure, therefore divided by 12 |
| >€288 but no logical explanation for being >€288 | Missing | 13 | No logical explanation for being >€288 |
| €1,250 | €12.50 | 2 | Medical card holder (€2.50 per medicine) and prescribed five regular medicines, therefore likely a mistaken decimal place |
| €750 | €7.50 | 3 | Medical card holder (€2.50 per medicine) and prescribed three regular medicines, therefore likely a mistaken decimal place |

### eTable 2. Characteristics of overall sample broken down by out-of-pocket (OOP) prescription medicine expenditure before scheme changes

|  | **Total % (n)** | **Mean OOP Prescription Medicine Expenditure (SD)** | **Median OOP Prescription Medicine Expenditure (IQR)** |
| --- | --- | --- | --- |
| **Age (years)** |  |  |  |
| <60 | 19.0 (1,075) | €254 (€439) | €60 (€0-300) |
| 60-69 | 40.9 (2,318) | €330 (€486) | €139.2 (€0-360) |
| 70-79 | 27.8 (1,574) | €325 (€464) | €153 (€60-300) |
| 80-89 | 11.2 (637) | €378 (€500) | €204 (€120-300) |
| 90+ | 1.1 (64) | €246 (€335) | €147 (€90-300) |
| **Sex** |  |  |  |
| Female | 55.6 (3,153) | €310 (€459) | €144 (€0-300) |
| Male | 44.4 (2,515) | €329 (€489) | €144 (€0-360) |
| **Education** |  |  |  |
| Primary/none | 23.6 (1,332) | €241 (€343) | €150 (€60-300) |
| Secondary | 39.6 (2,240) | €306 (€469) | €120 (€0-300) |
| Third/higher | 36.9 (2,096) | €381 (€535) | €156 (€0-480) |
| **Equivalised household income (quintiles)*** |  |  |  |
| <€6,000 | 21.3 (1,058) | €222 (€342) | €144 (€60-270) |
| €6,006-€10,000 | 18.9 (941) | €253 (€382) | €120 (€30-300) |
| €10,029-€15,133 | 19.9 (986) | €340 (€504) | €150 (€30-336) |
| €15,142-€24,000 | 20.6 (1,022) | €371 (€521) | €144 (€0-480) |
| >€24,000 | 19.3 (959) | €409 (€547) | €180 (€0-600) |
| **Area of residence*** |  |  |  |
| Urban | 52.0 (2,873) | €366 (€518) | €168 (€0-360) |
| Not Urban | 48.0 (2,648) | €270 (€415) | €120 (€0-300) |
| **Marital status** |  |  |  |
| Partnered | 69.0 (3,911) | €341 (€504) | €144 (€0-360) |
| Not Partnered | 31.0 (1,757) | €269 (€389) | €150 (€30-300) |
| **Private health insurance*** |  |  |  |
| Yes | 60.4 (3,425) | €401 (€540) | €180 (€0-480) |
| No | 39.6 (2,242) | €193 (€304) | €120 (€24-240) |
| **Long term illness (LTI) scheme** |  |  |  |
| Yes | 7.1 (404) | €307 (€489) | €120 (€12-300) |
| No | 92.9 (5,264) | €319 (€471) | €144 (€0-312) |
| **GMS & DPS*** |  |  |  |
| GMS Scheme | 46.5 (2,632) | €150 (€184) | €120 (€60-240) |
| Non-GMS (DPS eligible) | 53.5 (3,029) | €465 (€585) | €240 (€0-€720) |
| **Number of prescription medicines (quintiles)*^,+^** |  |  |  |
| 0 regular medicines^**^ | 25.2 (1,429) | €36 (€150)** | €0 (€0-0) |
| 1 regular medicine | 17.0 (965) | €235 (€362) | €120 (€30-240) |
| 2-3 regular medicines | 27.2 (1,539) | €389 (€457) | €240 (€90-480) |
| 4-5 regular medicines | 16.7 (948) | €514 (€554) | €240 (€144-720) |
| 6+ regular medicines | 13.9 (786) | €562 (€601) | €300 (€204-720) |
| **Number of chronic conditions** |  |  |  |
| 0 chronic conditions | 17.1 (967) | €111 (€300) | €0 (€0-72) |
| 1 chronic condition | 24.9 (1,411) | €251 (€410) | €120 (€0-300) |
| 2 chronic conditions | 23.4 (1,327) | €374 (€489) | €180 (€60-432) |
| 3+ chronic conditions | 34.6 (1,963) | €432 (€528) | €240 (€120-456) |
| **Complex multimorbidity** |  |  |  |
| Yes | 2.0 (114) | €360 (€433) | €240 (€120-300) |
| No | 98.0 (5,554) | €318 (€473) | €144 (€0-312) |

*These variables had data missing and the relevant percentages are calculated with missing data excluded.

^+^ Excluded medications not covered by state drug schemes as these are primarily over-the-counter medicines and supplements. Also excluded medications always indicated for a condition covered by the LTI scheme.

** Though these participants are on 0 regular medicines, the expenditure question does not preclude participants from including expenditure on non-regular medicines

Note: SD=Standard Deviation. IQR=Interquartile Range. GMS=General Medical Services. DPS=Drugs Payment Scheme

### eTable 3. Number (%) of people whose annual out-of-pocket prescription medicine expenditure is affected by change to DPS monthly limit, compared to previous threshold (e.g. €80 vs. €100)

|  | **€134 limit** | **€124 limit** | **€114 limit** | **€100 limit** | **€80 limit** |
| --- | --- | --- | --- | --- | --- |
| **Age (years)** |  |  |  |  |  |
| <60 | 14.8% (57) | 15.1% (58) | 16.1% (62) | 16.4% (63) | 20.1% (77) |
| 60-69 | 15.4% (148) | 16.2% (156) | 17.8% (171) | 18.3% (176) | 23.4% (225) |
| 70-79 | 22.2% (101) | 23.5% (107) | 26.2% (119) | 26.4% (120) | 32.5% (148) |
| 80+ | 30.7% (43) | 33.6% (47) | 35.7% (50) | 37.1% (52) | 51.4% (72) |
| **Sex** |  |  |  |  |  |
| Female | 17.2% (180) | 18.3% (191) | 19.5% (204) | 20.2% (211) | 25.8% (270) |
| Male | 18.9% (169) | 19.8% (177) | 22.1% (198) | 22.3% (200) | 28.2% (252) |
| **Education** |  |  |  |  |  |
| Primary/none | 18.5% (39) | 18.5% (39) | 20.9% (44) | 21.3% (45) | 26.5% (56) |
| Secondary | 18.8% (137) | 19.8% (144) | 21.2% (154) | 21.9% (159) | 28.2% (205) |
| Third/higher | 17.2% (173) | 18.4% (185) | 20.3% (204) | 20.6% (207) | 26.0% (261) |
| **Equivalised household income (quintiles)*** |  |  |  |  |  |
| <€6,000 | 17.0% (26) | 17.6% (27) | 19.0% (29) | 20.9% (32) | 26.8% (41) |
| €6,006-€10,000 | 15.8% (35) | 15.8% (35) | 16.2% (36) | 16.2% (36) | 22.1% (49) |
| €10,029-€15,133 | 21.0% (70) | 22.8% (76) | 24.9% (83) | 25.2% (84) | 32.1% (107) |
| €15,142-€24,000 | 18.8% (86) | 19.7% (90) | 21.2% (97) | 21.4% (98) | 27.8% (127) |
| >€24,000 | 17.0% (89) | 18.1% (95) | 20.0% (105) | 20.6% (108) | 25.5% (134) |
| **Area of residence** |  |  |  |  |  |
| Urban | 20.7% (228) | 22.0% (242) | 23.8% (262) | 24.5% (269) | 30.4% (334) |
| Not Urban* | 14.4% (121) | 15.0% (126) | 16.6% (140) | 16.9% (142) | 22.4% (188) |
| **Marital Status** |  |  |  |  |  |
| Married/cohabiting | 19.3% (289) | 20.3% (303) | 22.3% (333) | 22.6% (338) | 27.9% (418) |
| Not married/cohabiting | 13.5% (60) | 14.6% (65) | 15.5% (69) | 16.4% (73) | 23.4% (104) |
| **Private Health Insurance** |  |  |  |  |  |
| Yes | 18.7% (309) | 19.6% (325) | 21.6% (357) | 22.0% (364) | 27.8% (461) |
| No | 14.0% (40) | 15.1% (43) | 15.8% (45) | 16.5% (47) | 21.4% (61) |
| **Long Term Illness (LTI) Scheme** |  |  |  |  |  |
| Yes | 27.9% (29) | 28.8% (30) | 29.8% (31) | 30.8% (32) | 35.6% (37) |
| No | 17.4% (320) | 18.4% (338) | 20.2% (371) | 20.6% (379) | 26.4% (485) |
| **GP Entitlements** |  |  |  |  |  |
| GP Visit Card | 24.6% (124) | 25.7% (130) | 28.3% (143) | 28.7% (145) | 37.2% (188) |
| No GP Visit Card | 15.7% (225) | 16.6% (238) | 18.0% (259) | 18.5% (266) | 23.3% (334) |
| **Number of Prescription Medicines (quintiles)**** |  |  |  |  |  |
| 0 regular medicines | 5.1% (4) | 5.1% (4) | 6.3% (5) | 6.3% (5) | 10.1% (8) |
| 1 regular medicine | 5.7% (30) | 5.8% (31) | 6.2% (33) | 6.2% (33) | 8.3% (44) |
| 2-3 regular medicines | 11.1% (85) | 12.2% (93) | 14.1% (108) | 14.2% (109) | 19.6% (150) |
| 4-5 regular medicines | 31.8% (114) | 32.7% (117) | 35.5% (127) | 36.3% (130) | 44.7% (160) |
| 6+ regular medicines | 55.5% (116) | 58.9% (123) | 61.7% (129) | 64.1% (134) | 76.6% (160) |
| **Number of Chronic Conditions** |  |  |  |  |  |
| 0 chronic conditions | 12.3% (20) | 12.3% (20) | 13.0% (21) | 13.0% (21) | 16.7% (27) |
| 1 chronic condition | 10.3% (57) | 11.0% (61) | 12.3% (68) | 12.5% (69) | 16.1% (89) |
| 2 chronic conditions | 15.8% (90) | 16.7% (95) | 18.4% (105) | 18.9% (108) | 24.9% (142) |
| 3+ chronic conditions | 27.8% (182) | 29.3% (192) | 31.8% (208) | 32.5% (213) | 40.3% (264) |
| **Complex Multimorbidity** |  |  |  |  |  |
| Yes | 28.3% (154) | 29.8% (162) | 32.4% (176) | 32.9% (179) | 41.4% (225) |
| No | 14.0% (195) | 14.7% (206) | 16.2% (226) | 16.6% (232) | 21.3% (297) |

### eTable 4. Number (%) of people whose annual out-of-pocket prescription medicine expenditure is affected by change to GMS monthly threshold, compared to previous threshold (e.g. change 2 vs. change 1)

|  | **Change 1** | **Change 2** | **Change 3** |
| --- | --- | --- | --- |
| **Age (years)** |  |  |  |
| <60 | 94.1% (190) | 0.0% (0) | 94.1% (190) |
| 60-69 | 95.0% (606) | 0.0% (0) | 95.0% (606) |
| 70-79 | 96.0% (842) | 96.0% (842) | 96.0% (842) |
| 80+ | 94.6% (470) | 94.6% (470) | 94.6% (470) |
| **Sex** |  |  |  |
| Female | 96.4% (1255) | 57.3% (746) | 96.4% (1255) |
| Male | 93.5% (853) | 62.1% (566) | 93.5% (853) |
| **Education** |  |  |  |
| Primary/none | 95.0% (860) | 70.5% (638) | 95.0% (860) |
| Secondary | 95.2% (840) | 50.9% (449) | 95.2% (840) |
| Third/higher | 95.6% (408) | 52.7% (225) | 95.6% (408) |
| **Equivalised household income (quintiles)*** |  |  |  |
| <€6,000 | 95.1% (666) | 60.0% (420) | 95.1% (666) |
| €6,006-€10,000 | 95.5% (484) | 58.0% (294) | 95.5% (484) |
| €10,029-€15,133 | 95.2% (396) | 61.8% (257) | 95.2% (396) |
| €15,142-€24,000 | 95.1% (234) | 56.1% (138) | 95.1% (234) |
| >€24,000 | 98.0% (98) | 55.0% (55) | 98.0% (98) |
| **Area of residence** |  |  |  |
| Urban | 95.5% (1001) | 58.9% (617) | 95.5% (1001) |
| Not Urban* | 94.9% (1107) | 59.6% (695) | 94.9% (1107) |
| **Marital Status** |  |  |  |
| Married/cohabiting | 95.4% (1235) | 55.5% (718) | 95.4% (1235) |
| Not married/cohabiting | 94.9% (873) | 64.6% (594) | 94.9% (873) |
| **Private Health Insurance** |  |  |  |
| Yes | 96.1% (769) | 71.4% (571) | 96.1% (769) |
| No | 94.7% (1339) | 52.4% (741) | 94.7% (1339) |
| **Long Term Illness (LTI) Scheme** |  |  |  |
| Yes | 86.9% (173) | 42.7% (85) | 86.9% (173) |
| No | 96.0% (1935) | 60.9% (1227) | 96.0% (1935) |
| **Number of Prescription Medicines (quintiles)**** |  |  |  |
| 0 regular medicines | 0.0% (0) | 0.0% (0) | 0.0% (0) |
| 1 regular medicine | 100.0% (297) | 53.9% (160) | 100.0% (297) |
| 2-3 regular medicines | 100.0% (705) | 57.9% (408) | 100.0% (705) |
| 4-5 regular medicines | 100.0% (560) | 62.9% (352) | 100.0% (560) |
| 6+ regular medicines | 100.0% (546) | 71.8% (392) | 100.0% (546) |
| **Number of Chronic Conditions** |  |  |  |
| 0 chronic conditions | 90.8% (128) | 53.9% (76) | 90.8% (128) |
| 1 chronic condition | 95.1% (388) | 56.6% (231) | 95.1% (388) |
| 2 chronic conditions | 96.5% (526) | 58.7% (320) | 96.5% (526) |
| 3+ chronic conditions | 95.2% (1066) | 61.2% (685) | 95.2% (1066) |
| **Complex Multimorbidity** |  |  |  |
| Yes | 95.3% (918) | 60.9% (586) | 95.3% (918) |
| No | 95.1% (1190) | 58.0% (726) | 95.1% (1190) |

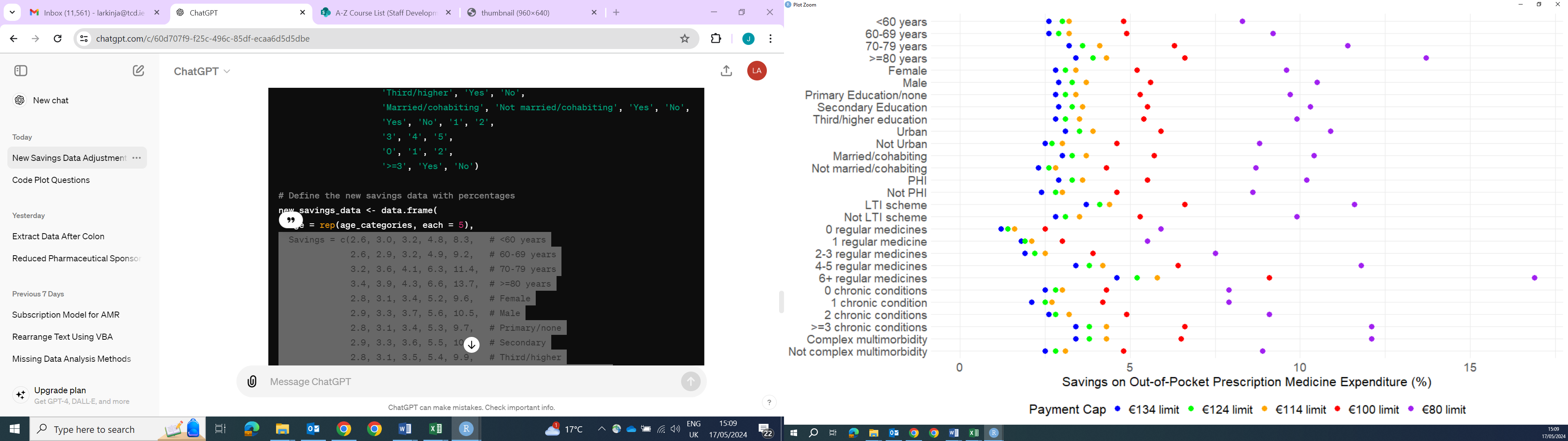

### eFigure 1. Mean percentage savings* for each DPS scheme limit, compared to previous limit (e.g. €80 vs. €100)

*Values for overlapping points have been slightly adjusted to improve visibility.

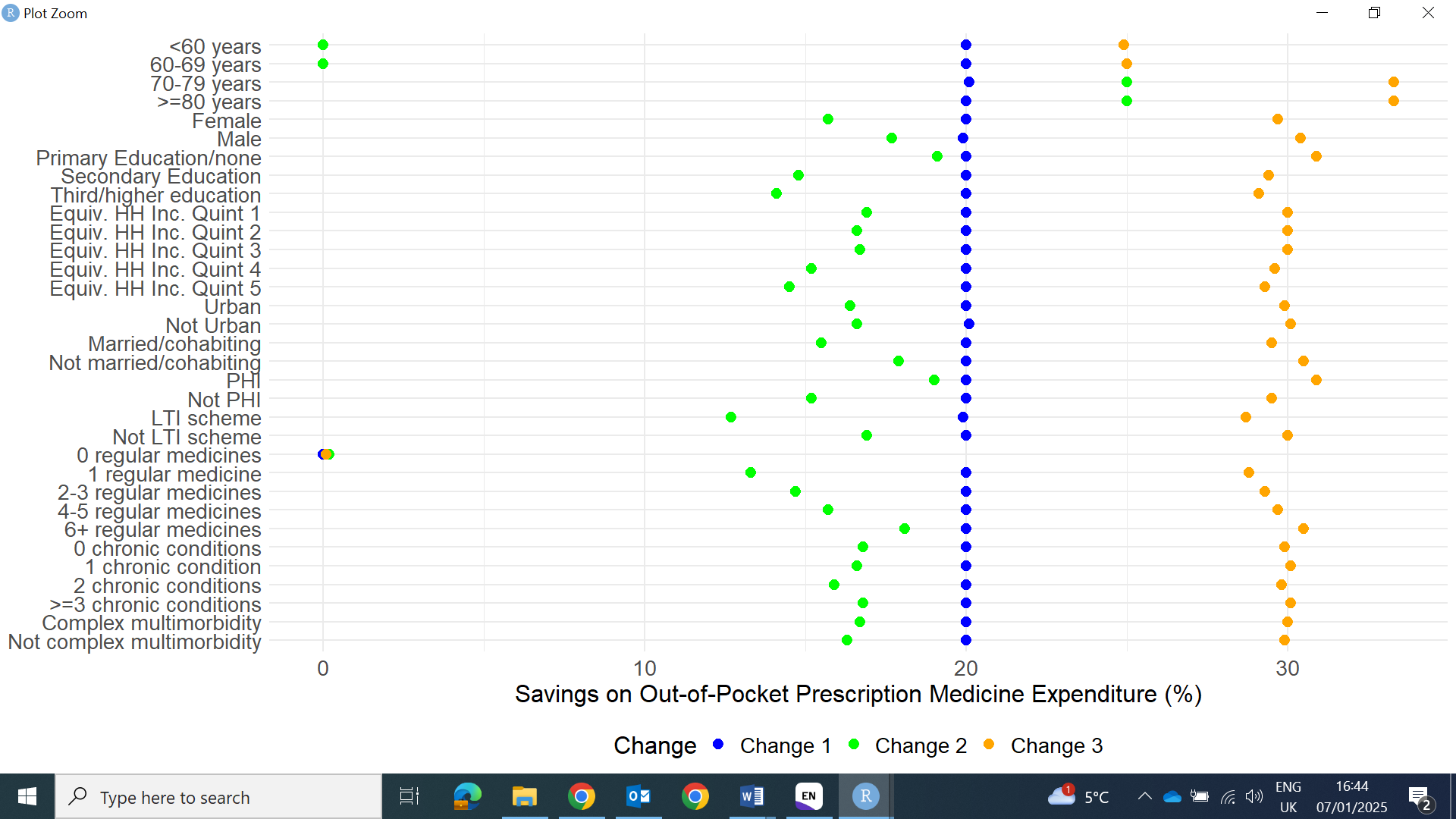
 eFigure 2. Mean percentage savings* for each scenario in the GMS group, compared to previous scenario (e.g. Change 3 vs. Change 2)

*Values for overlapping points have been slightly adjusted to improve visibility.
